# Location dependent weather condition is a factor that induces delirium requiring hospitalization in elderly patients: retrospective, single-center cohort study

**DOI:** 10.1101/2025.04.20.25326130

**Authors:** Yusuke Takatsuru, Noriaki Hattori

## Abstract

**Background:** Several factors for delirium have already been examined. However, the effects of location and weather conditions have not been fully evaluated yet. The aim of this study is to determine the effects of these factors on delirium in older adult patients.

**Methods:** We evaluated the data of older adult patients with and without delirium in the acute state on hospitalization from January 2017 to December 2020 in Johmoh Hospital. We also subdivided the patients into those who came from their own homes (COH group, 24 males and 20 females with delirium; 131 males and 218 females without delirium) and those who came from other places (COP group, 14 males and 16 females with delirium; 171 males and 284 females without delirium).

**Results:** We found that the number of delirious patients is higher in the COH group than the COP group. We also found that decreases in temperature, sunlight time, and average atmospheric pressure, and an increase in humidity were significant factors that induce delirium in COH group.

**Conclusion:** The risk factor for delirium differs depending on the housing condition, and weather is one of the important factors that should be considered to prevent delirium especially in COH group.

## Introduction

Clinicians often encounter cases of delirium in older adults, which is characterized by changes in the consciousness state, during general medical treatment. Several diseases induce delirium, and hospitalization itself is one of the main triggers of delirium.^1–3^ Delirious patients are difficult to keep calm, and they frequently make noises and remove their intravenous needles themselves. Moreover, there are patients with hypoactive delirium whose symptoms vary and sometimes difficult to distinguish from other diseases such as depression and apathy in dementia. These definitely disrupt acute and chronic medical care. A delay of treatment of underlying diseases also worsens delirium; thus, delirium is one of the most difficult conditions to deal with in general medical care. Therefore, the appropriate treatment of delirium is important especially in Japan because it has the largest super-aging population worldwide.

The treatment and care management of delirium are clearly described in several clinical guidelines.^4,5^ The guidelines stipulate that antipsychotic drugs such as haloperidol and quetiapine are sometimes required to treat delirium. However, general medical doctors not familiar with psychiatry hesitate to use these drugs because of the difficulty in choosing and controlling their doses. Moreover, the care management of delirium is sometimes difficult for general medical staff members in the general ward of a hospital. On the other hand, the treatment of physical diseases that induce delirium is sometimes difficult in a general psychiatric ward. Generally, it is easier to treat all older adult patients with delirium in specific hospitals that have both general and psychiatric departments.^2^ However, the number of beds in such hospitals is insufficient for the number of older adult patients who may potentially suffer from delirium. Thus, if we can identify predictors of delirium in older adults, it may be useful for preventing the overburdening of medical resources.

Weather conditions are strong candidates as predictors of the occurrence of delirium because, for example, dehydration is one of the important triggers of delirium.^1–3^ Japanese older adults tend to hesitate to use their air conditioner in summer and frequently overuse heating appliances such as electric blankets and foot warmers in winter. These home conditions and changes in weather conditions potentially induce delirium. However, the relationship between weather conditions and delirium has not been well studied. Moreover, housing conditions differ among older adults. One of the different between own home and other places such as acute-care hospitals and older-adult care facilities may the control of living condition depend on weather condition. Other places such as acute-care hospitals and older-adult care facilities potentially under the control of temperature and humidity because of the medical staff care about it using air conditioner and heating appliances. In contrast, living condition in own home is potentially out of control because of older patients control the air conditioner and heating appliances by themselves.

In this study, we observed patients aged over 65 years with and without delirium at the time of admission to our hospital and compared the patients coming from their own homes (COH group) and those coming from other places (COP group) such as acute-care hospitals and older-adult care facilities by retrospective cohort study manner. We also compared the weather conditions during the same period. Our results suggest that the number of patients who have delirium on hospitalization day was significantly higher in COH group. We also found that specific weather conditions increase the enhance of delirium occurring in COH group.

## Materials and Methods

### Ethical approval

This study was approved by the ethics committee of Toyo University (TU2020-09-TU2021-K003-TU2021-K005-TU2023-K049) and performed in accordance with the Declaration of Helsinki. This study was performed as retrospective, single center cohort study.^4,5^

### Background of patients

We evaluated the clinical records of patients aged over 65 years who were hospitalized from January 2017 to December 2020 in the acute-care ward (both psychiatry and internal medicine ward) of Johmoh Hospital (a psychiatric hospitals with an internal medicine ward), Japan. They were followed up after hospitalization until May 2024. We included all emergency patients aged over 65 years including those who presented with pain, hypoxia, fever, and others (see also, Table 1). We excluded those who 1) decided to enter the chronic care ward at the time of hospitalization and those who 2) came back to our hospital after going to another hospital owing to acute diseases such as bone fractures. We divided the patients into those with delirium (44 males and 30 females) and without delirium (349 males and 455 females) at the time of hospitalization. Most of the delirium patients showed the hyperactive pattern of delirium and only a few showed the hypoactive pattern. Thus, we did not divide them according to their delirium pattern in this study. We did not include those who newly showed the symptoms of delirium after hospitalization. Delirium was diagnosed on the basis of the DSM-V at the time of hospitalization by highly experienced psychiatrists or designated physicians of mental health. The diagnoses of other complications shown in Table 1 were based on the ICD-10 or DSM-V. Diagnoses of some complications were based on DSM-IV and we did not modify them in this study. We also subdivided the patients into those who came to the hospital from their own homes (COH group, 24 males and 14 females with delirium; 131 males and 171 females without delirium) and other places such as acute-care hospitals and older-adult care facilities (COP group, 20 males and 16 females with delirium; 218 males and 284 females without delirium).

**Table 1.**
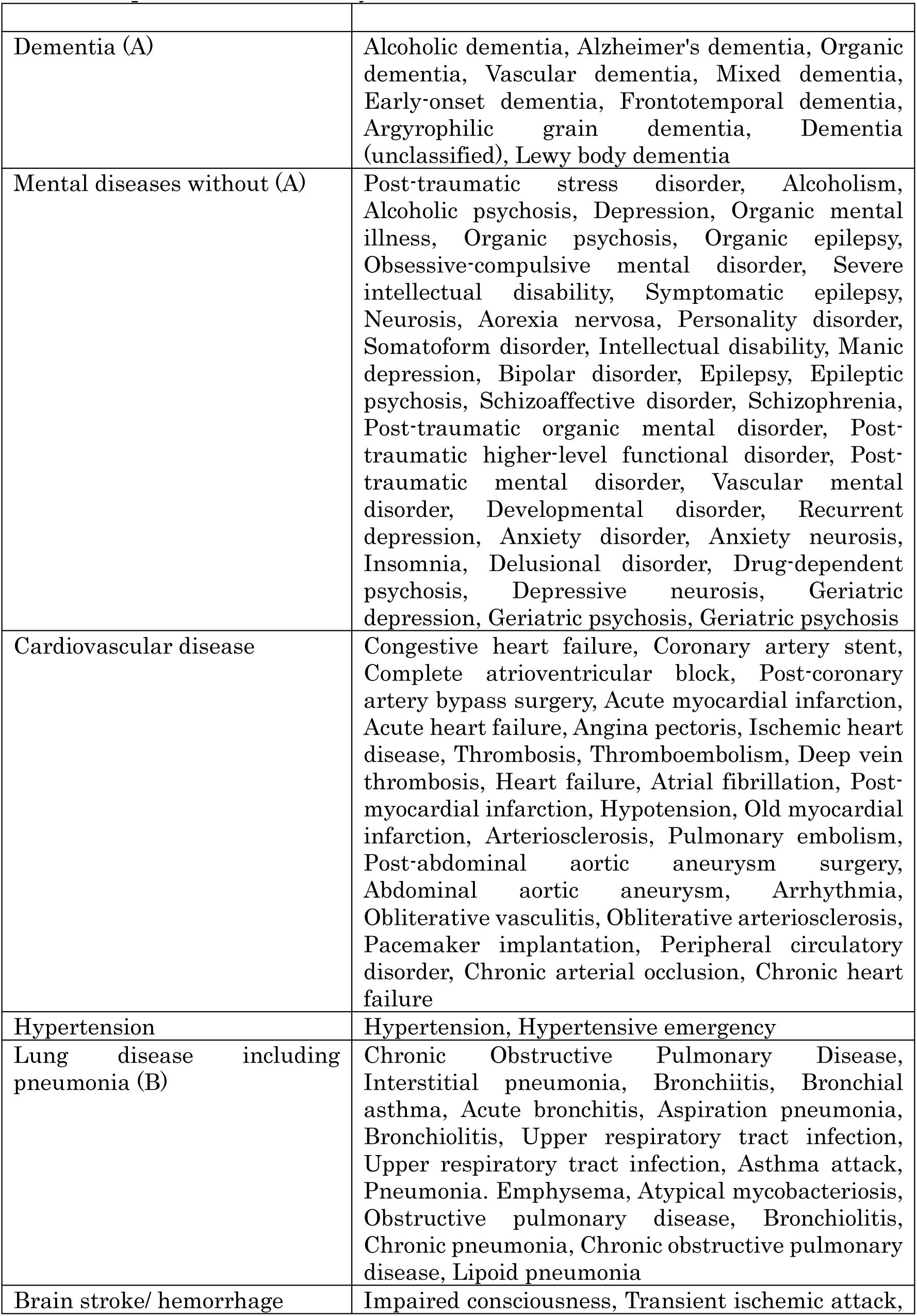
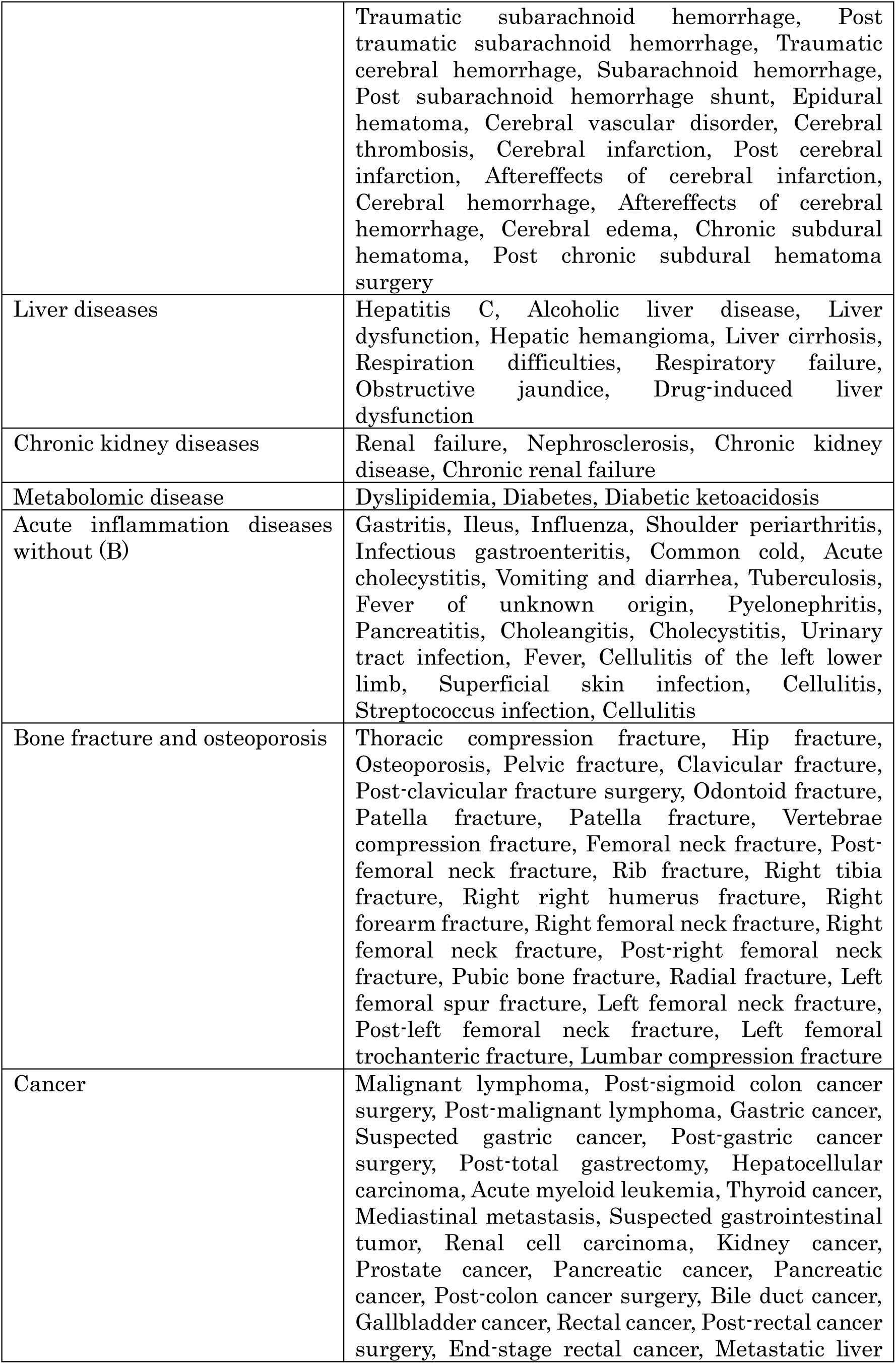
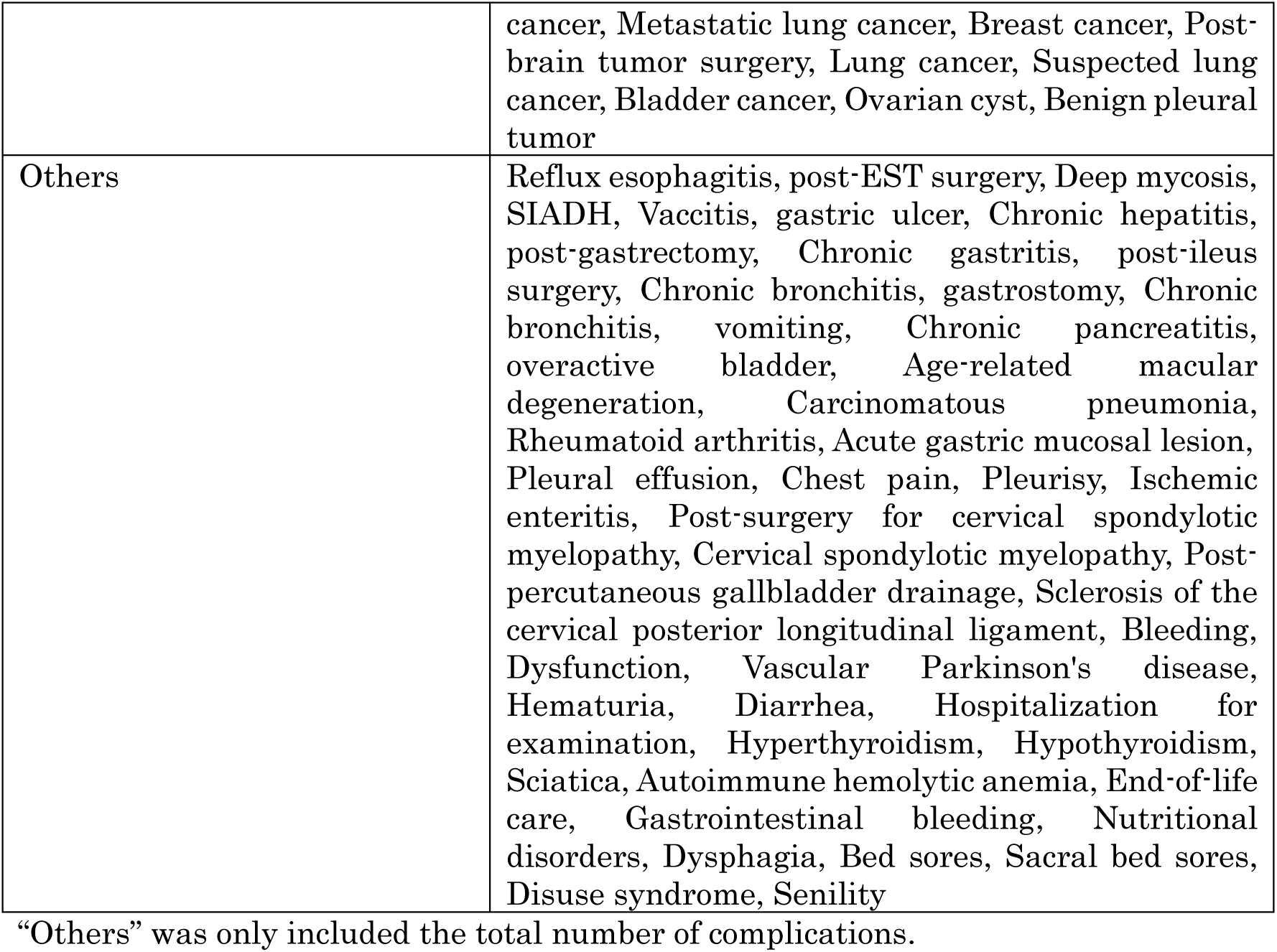
Complications in this study.

### Clinical and laboratory data

We determined the following at the time of hospitalization: the numbers of red blood cells (RBCs), white blood cells (WBCs), and platelets (Plt), the concentrations of hemoglobin (Hb), d-dimer, total protein (TP), albumin (Alb), aspartate aminotransferase (AST), alanine aminotransferase (ALT), γ-glutamyl transpeptidase (γGTP), creatine phosphokinase (CPK), blood urea nitrogen (BUN), creatinine (Cr), C-reactive protein (CRP), N-terminal pro-brain natriuretic peptide (NTProBNP), sodium (Na), potassium (K), and chloride (Cl), and the estimated glomerular filtration rate (eGFR) (Table 2). We also evaluated the complications and the drugs administered at the time of hospitalization (Tables 3 and 4). Some patients have several complications; and thus, the total number of complications was larger than the total number of patients. As shown in Table 3, we summarized the drugs that correlated with the delirium (both as risk factors and treatment agents).^6–8^

**Table 2.**
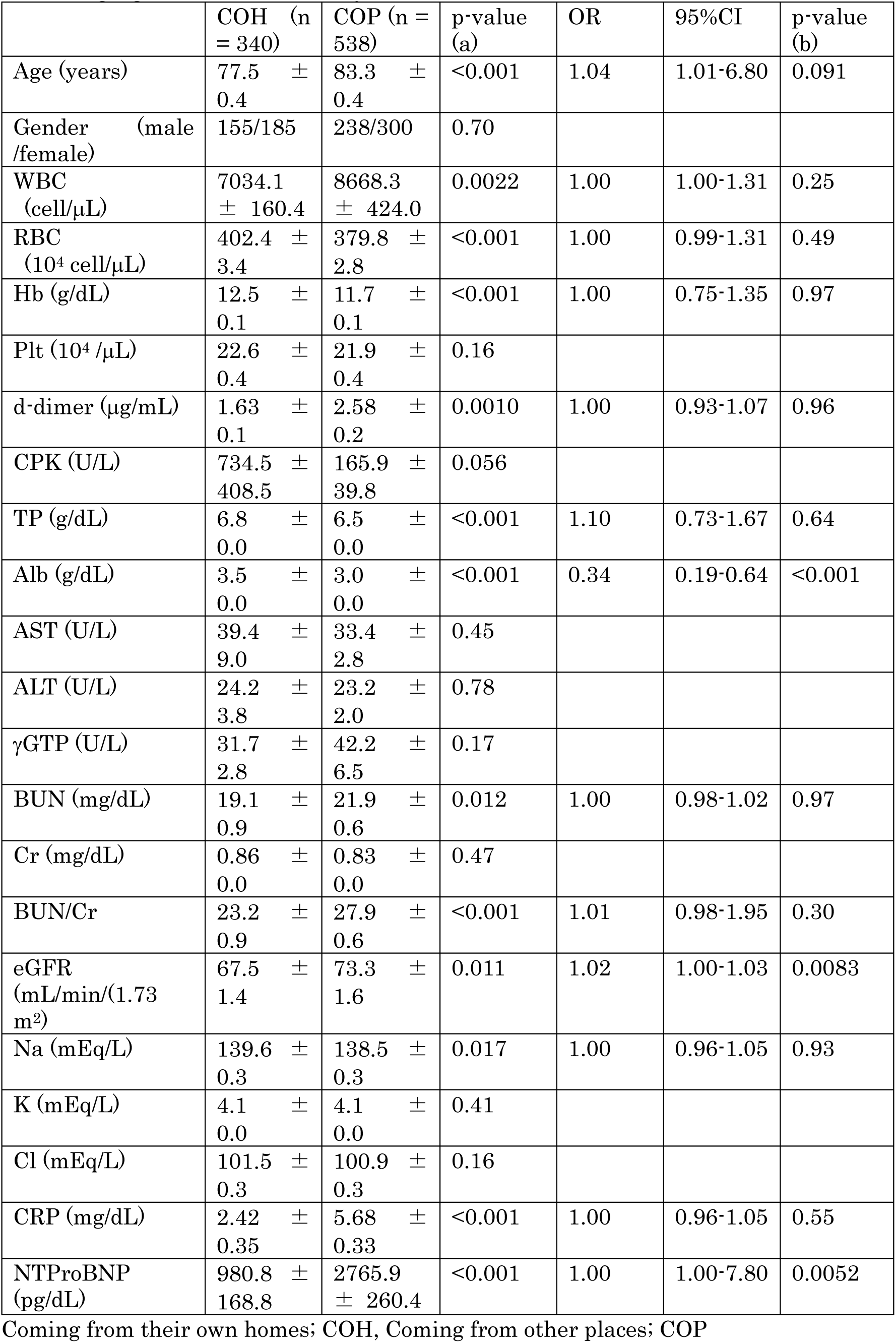
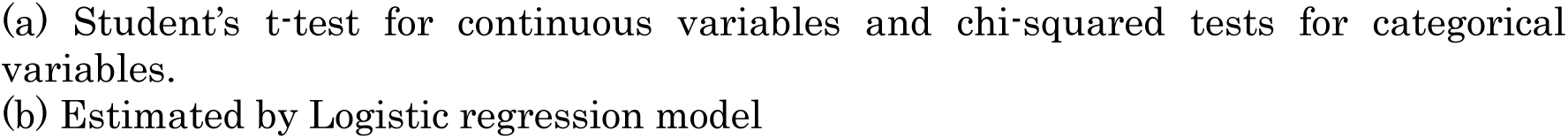
Age, gender and Laboratory data.

|  | COH (n = 340) | COP (n = 538) | p-value (a) | OR | 95%CI | p-value (b) |
| --- | --- | --- | --- | --- | --- | --- |
| Age (years) | 77.5 ± 0.4 | 83.3 ± 0.4 | <0.001 | 1.04 | 1.01-6.80 | 0.091 |
| Gender (male /female) | 155/185 | 238/300 | 0.70 |  |  |  |
| WBC (cell/μL) | 7034.1 ± 160.4 | 8668.3 ± 424.0 | 0.0022 | 1.00 | 1.00-1.31 | 0.25 |
| RBC ($10^4$ cell/μL) | 402.4 ± 3.4 | 379.8 ± 2.8 | <0.001 | 1.00 | 0.99-1.31 | 0.49 |
| Hb (g/dL) | 12.5 ± 0.1 | 11.7 ± 0.1 | <0.001 | 1.00 | 0.75-1.35 | 0.97 |
| Plt ($10^4$ /μL) | 22.6 ± 0.4 | 21.9 ± 0.4 | 0.16 | | | |
| d-dimer (μg/mL) | 1.63 ± 0.1 | 2.58 ± 0.2 | 0.0010 | 1.00 | 0.93-1.07 | 0.96 |
| CPK (U/L) | 734.5 ± 408.5 | 165.9 ± 39.8 | 0.056 |  |  |  |
| TP (g/dL) | 6.8 ± 0.0 | 6.5 ± 0.0 | <0.001 | 1.10 | 0.73-1.67 | 0.64 |
| Alb (g/dL) | 3.5 ± 0.0 | 3.0 ± 0.0 | <0.001 | 0.34 | 0.19-0.64 | <0.001 |
| AST (U/L) | 39.4 ± 9.0 | 33.4 ± 2.8 | 0.45 |  |  |  |
| ALT (U/L) | 24.2 ± 3.8 | 23.2 ± 2.0 | 0.78 |  |  |  |
| γGTP (U/L) | 31.7 ± 2.8 | 42.2 ± 6.5 | 0.17 | | | |
| BUN (mg/dL) | 19.1 ± 0.9 | 21.9 ± 0.6 | 0.012 | 1.00 | 0.98-1.02 | 0.97 |
| Cr (mg/dL) | 0.86 ± 0.0 | 0.83 ± 0.0 | 0.47 |  |  |  |
| BUN/Cr | 23.2 ± 0.9 | 27.9 ± 0.6 | <0.001 | 1.01 | 0.98-1.95 | 0.30 |
| eGFR (mL/min/(1.73 m<sup>2</sup>)) | 67.5 ± 1.4 | 73.3 ± 1.6 | 0.011 | 1.02 | 1.00-1.03 | 0.0083 |
| Na (mEq/L) | 139.6 ± 0.3 | 138.5 ± 0.3 | 0.017 | 1.00 | 0.96-1.05 | 0.93 |
| K (mEq/L) | 4.1 ± 0.0 | 4.1 ± 0.0 | 0.41 |  |  |  |
| Cl (mEq/L) | 101.5 ± 0.3 | 100.9 ± 0.3 | 0.16 |  |  |  |
| CRP (mg/dL) | 2.42 ± 0.35 | 5.68 ± 0.33 | <0.001 | 1.00 | 0.96-1.05 | 0.55 |
| NTProBNP (pg/dL) | 980.8 ± 168.8 | 2765.9 ± 260.4 | <0.001 | 1.00 | 1.00-7.80 | 0.0052 |

Coming from their own homes; COH, Coming from other places; COP
- (a) Student's t-test for continuous variables and chi-squared tests for categorical variables. - (b) Estimated by Logistic regression model

**Table 3.** Complications.

|  | COH (n = 340) | COP (n = 538) | p-value (a) | OR | 95%CI | p-value (b) |
| --- | --- | --- | --- | --- | --- | --- |
| Number of Complications | 3.6 ± 0.1 | 4.0 ± 0.1 | <0.001 | 1.27 | 1.10-1.47 | <0.001 |
| Dementia (A) | 97 | 175 | 0.21 |  |  |  |
| Mental diseases without (A) | 209 | 186 | <0.001 | 0.43 | 0.31-0.58 | <0.001 |
| Cardiovascular disease | 70 | 165 | 0.0010 | 1.17 | 0.80-1.71 | 0.43 |
| Hypertension | 181 | 223 | <0.001 | 0.42 | 0.29-0.60 | <0.001 |
| Lung disease including pneumonia (B) | 41 | 167 | <0.001 | 2.51 | 1.67-3.77 | <0.001 |
| Brain stroke/hemorrhage | 34 | 117 | <0.001 | 1.87 | 1.19-2.93 | 0.0063 |
| Liver diseases | 8 | 13 | 0.95 |  |  |  |
| Chronic kidney diseases | 142 | 226 | 0.94 |  |  |  |
| Metabolomic disease | 99 | 129 | 0.091 |  |  |  |
| Acute inflammation diseases without (B) | 17 | 106 | <0.001 | 4.17 | 2.39-7.27 | <0.001 |
| Bone fracture and osteoporosis | 39 | 68 | 0.61 |  |  |  |
| Cancer | 24 | 35 | 0.75 |  |  |  |

**Table 4.** Number of patients who were administrated drugs.

|  | COH (n = 340) | COP (n = 538) | p-value (a) | OR | 95%CI | p-value (b) |
| --- | --- | --- | --- | --- | --- | --- |
| Number of drugs per patients | 5.0 ± 0.2 | 4.9 ± 0.1 | 0.61 |  |  |  |
| Benzodiazepines/ benzodiazepine-like | 139 | 148 | <0.001 | 0.54 | 0.40-0.72 | <0.001 |
| Antiepileptic | 17 | 17 | 0.17 |  |  |  |
| NSAIDs | 11 | 14 | 0.58 |  |  |  |
| Antiparkinsonian | 15 | 29 | 0.52 |  |  |  |
| Antidepressants | 5 | 2 | 0.075 |  |  |  |
| Antipsychotic (without Quetiapine, Haloperidol, Risperdal) | 17 | 13 | 0.040 | 0.55 | 0.25-1.18 | 0.12 |
| Lithium carbonate | 14 | 17 | 0.45 |  |  |  |
| Antiarrhythmic | 6 | 8 | 0.75 |  |  |  |
| Digitalis | 4 | 10 | 0.43 |  |  |  |
| Antihypertensive | 115 | 224 | 0.021 | 1.45 | 1.09-1.93 | 0.012 |
| Xanthine | 6 | 7 | 0.58 |  |  |  |
| H2-blockers | 9 | 6 | 0.088 |  |  |  |
| Prednisolone | 5 | 9 | 0.81 |  |  |  |
| Antibiotics | 1 | 2 | 0.85 |  |  |  |
| Antihistamines | 11 | 5 | 0.70 |  |  |  |
| Quetiapine | 45 | 80 | 0.50 |  |  |  |
| Haloperidol | 3 | 12 | 0.13 |  |  |  |
| Risperdal | 12 | 37 | 0.035 | 2.39 | 1.21-4.71 | 0.012 |

NSAIDs; non-steroidal anti-inflammatory drugs

### Weather data

Weather data of Maebashi, Gunma, Japan, including the average, maximum, and minimum temperatures (Ave.T, Max.T, and Min.T, respectively), total sunlight time (TST), average atmospheric pressure (Ave.AP), and average humidity (Ave.H), were obtained from the home page of the Japan Meteorological Agency (https://www.jma.go.jp/jma/menu/menureport.html). Because changes in weather condition before hospitalization may induce delirium, we examined the weather data one day and two days before their hospitalization. Then, we determined ΔAve.T [Ave.T on (Day −1) – (Day −2)], ΔMax.T [Max.T on (Day −1) – (Day −2)], ΔMin.T [Min.T on (Day −1) – (Day −2)], ΔTST [TST on (Day −1) – (Day −2)], ΔAve.AP [Ave.AP on (Day −1) – (Day −2)], ΔAve.H [Ave.H on (Day −1) – (Day −2)] for statistical analysis as shown in Fig 1. If the value is below 0, it means that the value decreased from Day-2 to Day-1.

**Figure 1.**
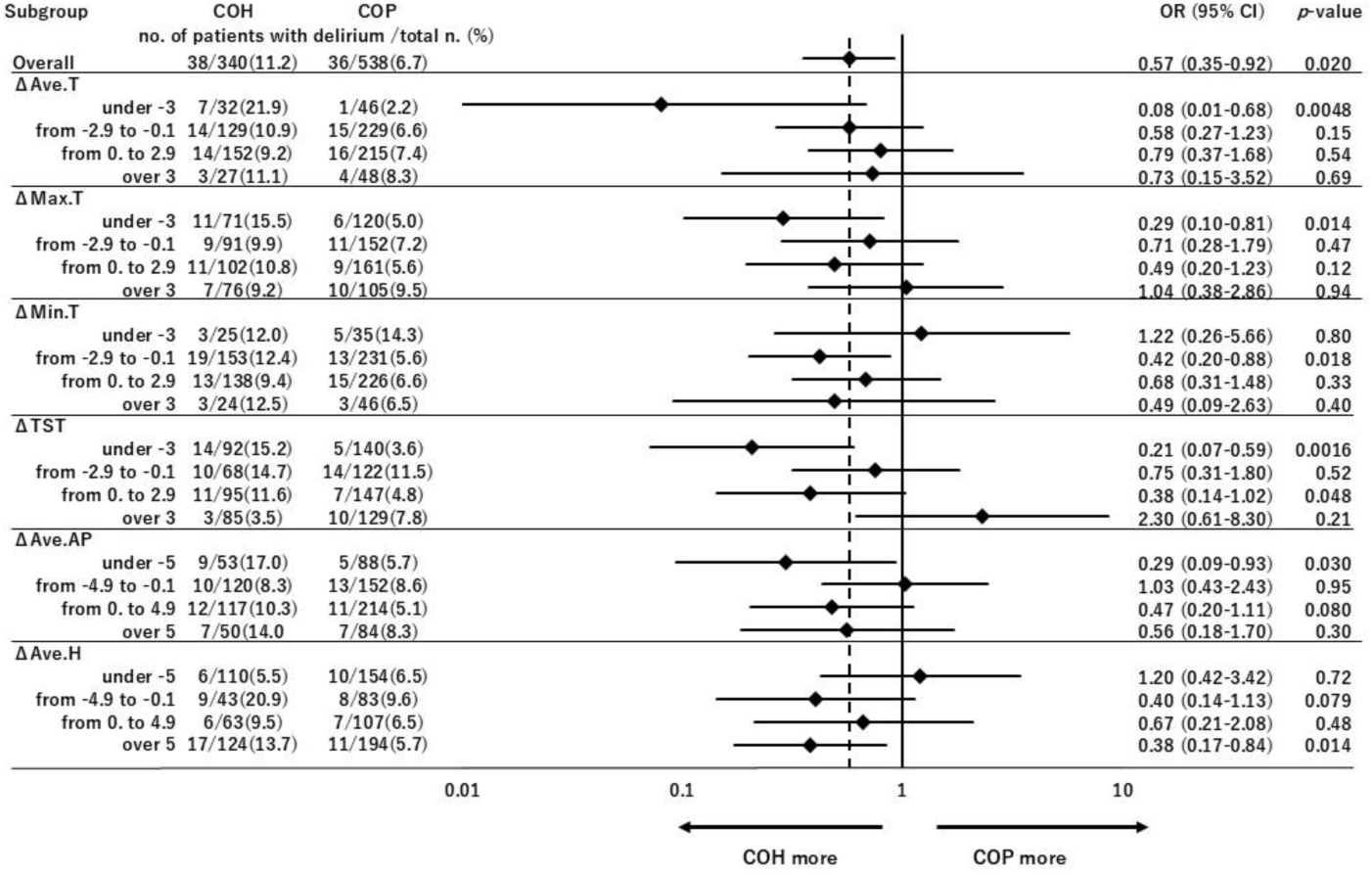
Changes in weather conditions are a risk factor for delirium in COH group. COH and COP groups were subdivided by weather condition and observed the percentage of the number of delirium patients in each group. ΔAve.T [Ave.T of (Day −1) – (Day −2)], ΔMax.T [Max.T of (Day −1) – (Day −2)], ΔMin.T [Min.T of (Day −1) – (Day −2)], ΔTST [TST of (Day −1) – (Day −2)], ΔAve.AP [Ave.AP of (Day −1) – (Day −2)], ΔAve.H [Ave.H of (Day −1) – (Day −2)]. OR and *p*-value was estimated by performed chi-squared tests.

### Statistical analyses

All the data were compared using Student’s *t*-test for continuous variables and chi-squared tests for categorical variables using BellCurve for Excel Ver4.07 (Social Survey Research Information Co., Ltd, Japan). Also, binary logistic regression analysis performed using BellCurve for Excel Ver4.07 (Social Survey Research Information Co., Ltd, Japan). We subdivided the groups depend on weather condition and compared the percentage of delirium patients in COH and COP.^9,10^ The difference was considered significant at *p* < 0.05. All values are presented as mean ± SEM.

## Results

In this study, we compared the effect of weather conditions on delirium at the time of hospitalization. Because of the use of air conditioners, the temperatures in the hospitals and older-adult care facilities are usually ideally constant and independent of the outside weather. Thus, we subdivided the patients into COH group and COP group.

As shown in Table 6, the number of delirium patients was significantly higher in COH group than in COP group (*p* = 0.020 by chi-squared tests). As shown in Tables 2, the Alb and NTProBNP concentrations and eGFR were significantly associated with the place where the patients came from. As shown in Table 3, the number of complications a patient had delirium, mental diseases without dementia, hypertension, lung diseases including pneumonia, brain stroke/ hemorrhage, and acute inflammation diseases without lung disease were significantly associated with the place where the patients came from. As shown in Table 4, benzodiazepines/benzodiazepine-like and antihypertensive drugs, and risperdal were significantly associated with the place where the patients came from. As shown in Table 16, we used binary logistic regression to control for those factors as potentially confounding roles and found that place of patients before hospitalization was significantly affects the induction of delirium.

**Table 6.**
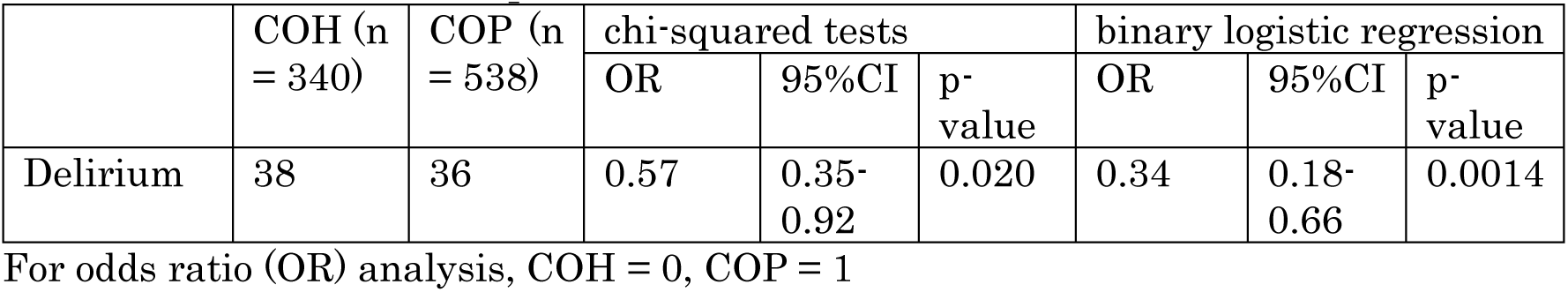
Number of delirium patients in COH and COP.

|  | COH (n<br>= 340) | COP (n<br>= 538) | chi-squared tests |  |  | binary logistic regression |  |  |
| --- | --- | --- | --- | --- | --- | --- | --- | --- |
|  |  |  | OR | 95%CI | p-value | OR | 95%CI | p-value |
| Delirium | 38 | 36 | 0.57 | 0.35-0.92 | 0.020 | 0.34 | 0.18-0.66 | 0.0014 |

For odds ratio (OR) analysis, COH = 0, COP = 1

We hypotheses that small change of weather condition may not affect the number of delirium patients and it may hide the difference between COH and COP groups if we using overall multi logistic analysis. Actually, binary logistic analysis not shown the statistical significance between housing condition and weather conditions (Table 7). Thus, as shown in Fig.1, we performed the stratified analysis for each weather conditions and compared the number of delirium patients both in COH and COP groups.

**Table 7.** Binary logistic analysis of delirium and weather conditions.

|  | OR | 95%CI | p-value (b) |
| --- | --- | --- | --- |
| Delirium<br>(Yes = 1, No = 0) | 0.58 | 0.36-0.94 | 0.027 |
| ΔAve.T | 1.06 | 0.85-1.31 | 0.60 |
| ΔMax.T | 0.97 | 0.87-1.07 | 0.54 |
| ΔMin.T | 1.01 | 0.89-1.14 | 0.90 |
| ΔTST | 0.99 | 0.95-1.04 | 0.77 |
| ΔAve.AP | 1.01 | 0.98-1.04 | 0.43 |
| ΔAve.H | 1.00 | 0.98-1.02 | 0.82 |

ΔAve.T [Ave.T of (Day -1) – (Day -2)], ΔMax.T [Max.T of (Day -1) – (Day -2)], ΔMin.T [Min.T of (Day -1) – (Day -2)], ΔTST [TST of (Day -1) – (Day -2)], ΔAve.AP [Ave.AP of (Day -1) – (Day -2)], ΔAve.H [Ave.H of (Day -1) – (Day -2)]. COH = 0, COP = 1

We found that decreases in ΔAve.T (under −3 ℃, *p* = 0.0048), ΔMax.T (under −3 ℃, *p* = 0.014), ΔMin.T (from –2.9 to −0.1 ℃, *p* = 0.018), ΔTST (under −3 hr, *p* = 0.0016), and ΔAve.AP (under −5 mmHg, *p* = 0.030) induced delirium more in COH group than those in COP group. Moreover, increase in ΔMin.T (from –2.9 to −0.1 ℃, *p* = 0.018), ΔTST (under −3 hr, *p* = 0.0016. from 0 to 2.9 hr, *p* = 0.048) and ΔAve.H (over 5%, *p* = 0.014) also induced delirium more in COH group than those in COP group.

In conclusion, living conditions may induce delirium, and weather conditions are among the triggers of delirium in COH group. If we consider the results of our study, we may be able to predict the onset of delirium and take countermeasures by predicting the weather.

## Discussion

In this study, we analyzed the factors that trigger delirium requiring hospitalization in older adult patients and found that the housing and weather conditions induce delirium.

Weather conditions definitely affect human health.^11–14^ For example, a low air pressure was previously reported to induce several health conditions, such as headaches^15^ and mental disorders.^16^ It was reported that a decrease in air pressure was not correlated with headaches on the day of their onset but on the day after their onset.^15^ High temperatures may induce delirium,^17^ and changes in weather conditions with the change in season from late summer to early autumn may also induce delirium. ^18,19^ In this study, decreases in temperature and sunlight time, and an increase in humidity are risk factors for delirium especially in COH group. These factors are well controlled in acute-care hospitals and older-adult care facilities compared with those in private homes. Interestingly, small decrease (but not the big change) of ΔMin.T affected the number of delirium patients. Too much low Min.T may force older patients to control the housing condition (using air conditioner or heating appliances) and because of the reason, number of deliriums may not significantly different between COH and COP groups under low Min.T condition. Moreover, decrease in AP (which may not control both in COH and COP groups) and increase in TST also induces delirium. Further study of these finding is needed; weather conditions could be one of the risk factors for delirium in patients who live in their own homes.

One limitation of this study is that the change in temperature was analyzed using data from the Japan Meteorological Agency and not the actual temperature of each place. Moreover, the mechanisms underlying the relationship between temperature and delirium in females staying in their own homes are still not fully understood; therefore, the next step is to conduct a prospective study. Another limitation is that this was a single-center observational study, and the selection of the population of patients was potentially biased. A multicenter observation will be needed in the future.

Results of our study suggests that the patients staying in their own homes are potentially at risk of delirium because housing conditions are not well controlled with the changes in weather conditions. The prevention and rapid treatment of delirium are important for a stable medical system, and this study partly contributes to the medical field in terms of the prevention and treatment of delirium.

## Data Availability

All data produced in the present study are available upon reasonable request to the authors

## Acknowledgements

The authors have no potential acknowledgements to disclose.

## Notes

### Competing Interest Statement

The authors have declared no competing interest.

### Funding Statement

This study did not receive any funding

### Author Declarations

This study was approved by the ethics committee of Toyo University (TU2020-09-TU2021-K003-TU2021-K005-TU2023-K049) and performed in accordance with the Declaration of Helsinki.

